# Beyond Traditional Assessments of Cognitive Status: Exploring the Potential of Spatial Navigation Tasks

**DOI:** 10.1101/2024.10.12.24315402

**Authors:** Giorgio Colombo, Karolina Minta, Tyler Thrash, Jascha Grübel, Jan Wiener, Marios Avraamides, William R. Taylor, Christoph Hölscher, Victor R. Schinazi

## Abstract

Deficits in spatial and navigation abilities are among the earliest signs of dementia. Yet, traditional neuropsychological tests primarily target memory and attention. The Spatial Performance Assessment for Cognitive Evaluation (SPACE) is a novel gamified digital assessment for iPads that uses various spatial tasks to detect early deficits in spatial navigation abilities indicative of cognitive impairment. In this study, 348 participants aged 21–76 completed the Montreal Cognitive Assessment (MoCA), SPACE, and a sociodemographic and health questionnaire. We investigated whether SPACE could predict scores on the MoCA beyond known risk factors for cognitive impairment. Using a factor analysis, we then assessed whether SPACE could complement the MoCA by capturing latent variables independent of MoCA scores that represent additional spatial aspects of cognitive functioning. Results from a hierarchical regression revealed that the pointing and perspective taking tasks in SPACE significantly predicted MoCA scores beyond age and gender. Surprisingly, none of the risk factors predicted MoCA scores. The factor analysis revealed that the MoCA and perspective taking contributed to a separate factor from other navigation tasks in SPACE. We further provide normative data for age and gender for each task in SPACE, which can serve as benchmarks in future studies to identify individuals at risk.

## 1. INTRODUCTION

Dementia affects 55 million people worldwide (*World Health Organization*, 2023), and this number is projected to increase to 152 million by 2050, posing a significant global threat to the healthcare system (Patterson, 2018). Alzheimer’s Disease (AD) is the most common form of dementia and ranks as the fifth leading cause of death for those aged 65 and older (National Center for Health Statistics, 2021). The total economic burden of AD is projected to reach $3.3 trillion by 2060 (Nandi et al., 2024) and includes the costs of healthcare, long-term care, and informal caregiving (Alzheimer’s Association, 2024; Rajan et al., 2021). The early stage of dementia, known as Mild Cognitive Impairment (MCI), often goes undiagnosed due to its mild symptoms and gradual onset. This stage presents a critical window for deploying sensitive assessments that may help reduce economic and societal costs via early intervention. While researchers have identified several risk factors associated with MCI and AD (Livingston et al., 2020), other predictors, such as spatial navigation performance, may complement existing assessments.

In addition to unmodifiable risk factors such as age (*Alzheimer’s Disease Fact Sheet*, 2023; Hebert et al., 2010), APOE genetic status (Kunz et al., 2015), and family history (Alzheimer’s Association, 2024; Hebert et al., 2013), the Lancet Commission has identified 14 modifiable risk factors (e.g., education, depression, physical inactivity) that account for nearly half of the dementia cases globally (Livingston et al., 2024). Previous research has found that these factors and other related risks are associated with the outcomes of widely used clinical assessments for cognitive status, including the Montreal Cognitive Assessment (MoCA; Bugallo-Carrera et al., 2024; Dale et al., 2018; Del Brutto et al., 2015; Dupuis et al., 2015; Freire et al., 2017; Jia et al., 2021; Nasreddine et al., 2005; Zawar et al., 2022) and the Mini-Mental State Examination (MMSE; Folstein, 1983; Heymann et al., 2016; Jia et al., 2021; Kim, 2022; Lv et al., 2024; Paterniti et al., 2002; Xu et al., 2009). Specifically, worse performance on clinical assessments is associated with older age (Bugallo-Carrera et al., 2024; Dale et al., 2018; Jia et al., 2021), being female (Bugallo-Carrera et al., 2024; Jia et al., 2021), but see (Dale et al., 2018), less education (Bugallo-Carrera et al., 2024; Dale et al., 2018), worse health status (Dale et al., 2018), physical inactivity (Iso-Markku et al., 2024), higher depression (Bugallo-Carrera et al., 2024; Byers & Yaffe, 2011; Dale et al., 2018; Del Brutto et al., 2015), more anxiety and stress (Del Brutto et al., 2015; Potvin et al., 2011), history of alcohol consumption (Heymann et al., 2016; Xu et al., 2009), smoking (Jia et al., 2021), sleep (McSorley et al., 2019; Zawar et al., 2022), and lower hearing or vision (Dupuis et al., 2015). The MoCA is often preferred to the MMSE because of its greater sensitivity in the detection of MCI and AD (Jia et al., 2021; Julayanont & Nasreddine, 2017; Pinto et al., 2019; Tsai et al., 2016) and, in clinical practice, provides a cheaper and less invasive alternative to a full neuropsychological assessment and the measurement of biomarkers of neurodegeneration.

Critically, biomarkers such as tau and β-amyloid (Aβ) are known to accumulate in brain regions that are essential for spatial navigation (Fyhn et al., 2004; Weisberg et al., 2014), especially the hippocampus and entorhinal cortex (Barthélemy et al., 2020; Chételat et al., 2010; Jack et al., 2010; Jack & Holtzman, 2013; Schmidt-Hieber & Häusser, 2013). Specifically, place cells in the hippocampus and grid cells in the entorhinal cortex have been found to be key components in encoding spatial locations and tracking positional changes during navigation (Epstein et al., 2017; Hafting et al., 2005; McNaughton et al., 2006; O’Keefe & Nadel, 1979). These findings suggest that spatial ability may be an important predictor of future cognitive status. Indeed, previous research has shown that spatial abilities are among the first skills to deteriorate as a consequence of AD (Castegnaro et al., 2023; Coughlan et al., 2018; deIpolyi et al., 2007; Hort et al., 2007; Howett et al., 2019; Segen et al., 2022). Although the MoCA and neuropsychological assessments emphasise working memory and executive functions and include some basic visuospatial tasks, a more comprehensive assessment of spatial and navigation abilities may further improve the sensitivity of these assessments. Notably, visuospatial tasks often focus on the relations among items at the micro-scale and therefore tap only on a subset of the skills required to navigate a large environment. Navigation in large environments requires additional skills such as the apprehension and integration of spatial information from multiple viewpoints and awareness of one’s own movement through space (Hegarty et al., 2006; Meneghetti et al., 2021). Here, researchers have used several spatial tasks to predict MCI and AD with varying degrees of success (Berron, Glanz, et al., 2024; Chan et al., 2016; Coughlan et al., 2019; Hort et al., 2007; Rekers & Finke, 2024; Tu et al., 2015; van der Ham et al., 2020; Wiener et al., 2020) but have not systematically investigated the manner in which these spatial tasks can contribute to common cognitive assessments.

In the present study, we explore how spatial navigation performance, as measured by the Spatial Performance Assessment for Cognitive Evaluation (SPACE), relates to general cognitive functioning across the adult lifespan. SPACE is a novel gamified digital assessment that combines a variety of spatial tasks from previous research and is designed to identify deficits in spatial and navigation abilities indicative of early signs of cognitive impairment (Colombo et al., 2024). We hypothesise that known modifiable risk factors for dementia and performance in SPACE will be associated with general cognitive abilities assessed by the MoCA. In contrast, large-scale navigation tasks in SPACE may reflect additional spatial dimensions of cognitive function not captured by the MoCA. We also examine the influence of demographic factors such as age and gender on the various tasks in SPACE and aim to provide normative data across these variables to establish a foundation for future studies.

## 2. METHODS

### Participants

We collected data from 348 healthy participants between 21 and 76 years of age (M = 45, SD = 16) who were recruited via social media platforms (e.g., Facebook, LinkedIn, Telegram) and community flyers. This sample reflects the age distribution of individuals who were comfortable travelling independently to the Singapore-ETH Centre. Most of the recruited participants were from Singapore (n = 242), with the sample predominantly based in Asia overall (n = 273). Only small subsets were from Europe (n = 14), North America (n = 4), and South America (n = 1). Nationality data are missing for fifty participants. Individuals with neurological disorders, severe visual impairment or blindness, deafness, a history of seizures, epilepsy, or recent acute cardiac events were excluded from the study. Due to unforeseen technical issues, such as app crashes or refusal to answer questionnaires, six participants were entirely excluded from all analyses. Additionally, some participants had incomplete data entries, leading to missing values. These incomplete data were omitted from specific analyses, but the participants themselves were not completely excluded. Ultimately, data from 342 participants were included in the final analyses. A subset of this sample overlapped with an earlier SPACE usability study in which different control interfaces (Tap and Anchor conditions), a UI widget (Widget condition), and a simplified configuration of trials and landmarks (Simplified condition) were tested (Colombo et al., 2024). For the present study, all participants who completed the full SPACE protocol were pooled across the testing conditions from the usability study. Figure 1 shows the age distribution and the number of participants who scored < 26 in the MoCA.

**Figure 1.**
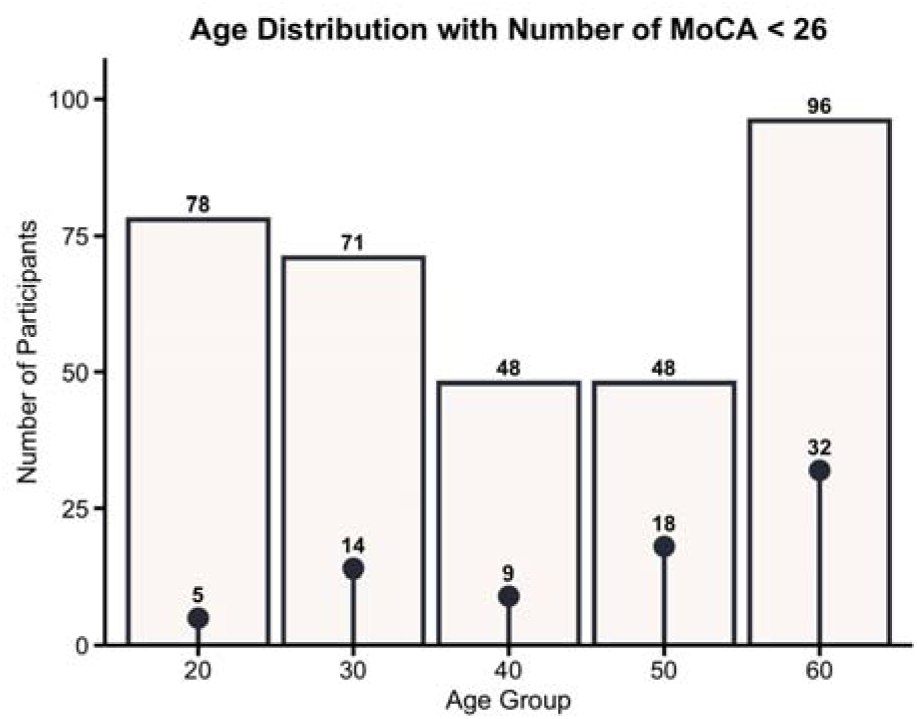
Age distribution and number of participants with MoCA scores < 26. Bar heights indicate the number of participants in each age group, and the overlaid points show the number of individuals scoring below the MoCA threshold for possible cognitive impairment.

Ethical approval for this study was granted by the Parkway Independent Ethics Committee (PIEC/2022/010) and the ETH Zurich Ethics Commission (EK 2021-N-193). Written informed consent was obtained from all participants prior to their involvement in the study. All procedures were conducted in accordance with the Declaration of Helsinki.

### Materials

#### Instruments

Participants completed both the MoCA and SPACE assessments. Before and after these assessments, participants were asked to complete a series of questionnaires, including a digital visual acuity test, a sociodemographic and health questionnaire, the Santa Barbara Sense of Direction scale (Hegarty et al., 2002), a series of usability questionnaires, and a debriefing questionnaire. The digital visual acuity test was based on a Snellen chart and was only used to screen participants for extreme visual impairments or blindness. The results from the usability and debriefing questionnaires are detailed in a separate paper (Colombo et al., 2024).

*MoCA*. The MoCA is a widely used cognitive screening tool designed to detect cognitive impairment. The MoCA is composed of a 30-point scale administered in person by a qualified examiner to assess various cognitive domains, including memory, executive function, visuospatial skills, language, attention, and orientation. A score below 26 typically indicates MCI. The MoCA has a sensitivity of 90% and a specificity of 87% for predicting cognitive impairment (Nasreddine et al., 2005). In addition, the MoCA (AUC values ranging from 0.71 to 0.99) has better diagnostic accuracy for MCI compared to the MMSE (AUC values ranging from 0.43 to 0.94; Pinto et al., 2019). For detecting AD, the MoCA also outperforms the MMSE, with AUC values ranging from 0.87 to 0.99 for the MoCA and 0.67 to 0.99 for the MMSE.

*SPACE*. SPACE is a novel gamified digital assessment designed to detect deficits in spatial navigation performance that may indicate signs of cognitive impairment (Colombo et al., 2024). SPACE is deployed on iPads and includes visuomotor training and five other spatial and navigation tasks. The visuomotor training is critical to minimise learning effects and ensure that performance does not reflect the ability to control the device (Grübel et al., 2017). In SPACE, participants navigate from a first-person perspective from one landmark to another to learn their relative positions as part of a path integration task. Participants are later probed on their spatial knowledge via pointing (Figure 2a), mapping, and associative memory tasks. Participants are also asked to complete a perspective taking task in which they are provided with a top-down representation of the landmarks (Figure 2b). These tasks are specifically developed to probe the acquisition of spatial knowledge at the environmental scale and are described in Table 1. The virtual environment is intentionally devoid of distinctive features apart from background mountains and the robot that guides participants to the landmarks. Participants encounter only two landmarks per trial and are unable to see a landmark when standing in front of another landmark. The landmarks only become visible when the participant approaches them and fade out when the participant moves away.

**Figure 2.**
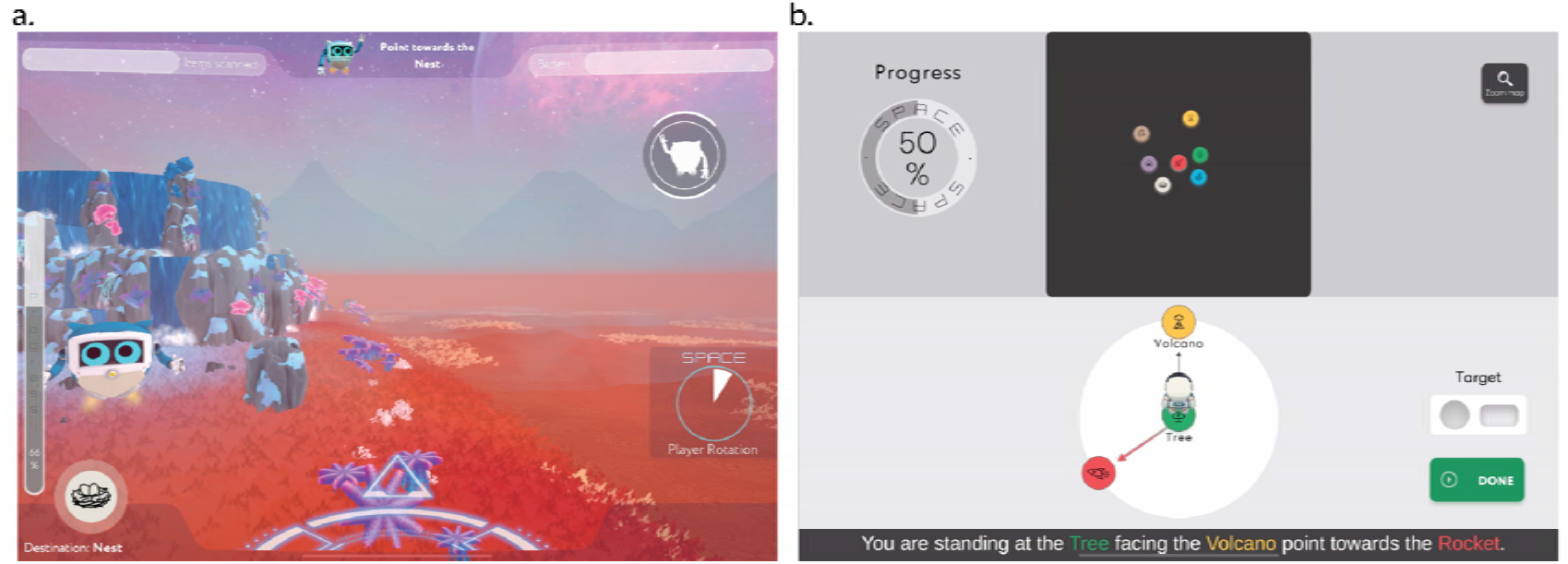
Screenshots from the pointing and perspective taking tasks in SPACE. (a) In the pointing task, participants are positioned at a landmark and asked to point toward another landmark encountered during the path integration task. (b) In the perspective taking task, participants imagine standing at one landmark and facing another. They must then adjust the target icon to indicate the correct direction to a third landmark from that perspective.

**Table 1.** Descriptions of the tasks in SPACE.

|  |  |
| --- | --- |
| **Visuomotor training** | Participants learn to rotate, translate, and integrate these movements by following a robot around the planet from a first-person perspective. In the final training phase, participants are introduced to the logic of the path integration task. |
| **Path integration** | Participants follow the robot from the rocket to two landmarks, walking along two sides of a triangle from a first-person perspective. At each landmark, the robot scans a different element that will be recalled in a later task. Participants are asked to return unguided to the original position of the rocket, completing the third side of the triangle. Unlike the final training phase, the rocket takes off at the start of each trial and stays invisible until participants signal its landing after completing the trial. |
| **Pointing** | Participants stand in front of a landmark or the rocket and are asked to complete a series of pointing trials to other landmarks encountered during the path integration task. |
| **Mapping** | Participants are asked to recreate the configuration of landmarks in the environment they learned by dragging and dropping icons representing the landmarks from a top-down perspective. |
| **Associative memory** | Participants are presented with a corrected top-down map of the landmarks and are asked to drag and drop icons representing the corresponding elements scanned by the robot during the path integration task. |
| **Perspective taking** | Participants are provided with the correct top-down map of the environment and are asked to imagine standing at a landmark while facing another landmark. Their task is to indicate the correct bearing toward a third landmark from this perspective. |

The different experimental conditions (i.e., Tap, Anchor, Widget, and Simplified) have different trial designs for each task. Path integration includes 13 trials in the Tap, Anchor, and Widget conditions, and 7 trials in the Simplified condition. The landmark set encountered during path integration forms the basis for all subsequent tasks. The egocentric pointing task includes 15 trials (Tap, Anchor,

Widget) or 9 trials (Simplified). In the mapping task and associative memory tasks, participants reconstruct the spatial configuration and landmark-item associations of all previously visited landmarks. The perspective taking task includes 13 trials (Tap, Anchor, Widget) or 7 trials (Simplified). A full top-down view of the environment and landmarks, together with breakdown of the number of participants assigned to each condition, is provided in Supplementary Information A.

*Sociodemographic and health questionnaire*. The sociodemographic and health questionnaire gathered information on the age, gender, education, background, handedness, tablet experience, and prior navigation training of participants. This questionnaire also collected data on their health status, including vision impairments, chronic conditions, and history of traumatic brain injury, as well as their psychosocial well-being, focusing on levels of depression, anxiety, and stress over the past six months. Additionally, the questionnaire addressed health habits, such as smoking, alcohol consumption, incidences of falls in the past year, daily hours of sleep, and weekly hours of walking and vigorous physical activity.

#### Hardware and Software

SPACE was deployed on a 10.2-inch iPad with Wi-Fi and 256 GB memory running iOS version 16.6.1. The vision test was conducted using the iPad app MDCalc (https://www.mdcalc.com). All questionnaire data were collected via the Qualtrics XM online survey platform (www.qualtrics.com) on the iPad. Gait data was collected using WitMotion sensors (WT901BLECL Bluetooth 5.0 Accelerometer, https://www.wit-motion.com).

### Procedure

Before starting the experiment, the experimenter briefed participants on the aim of the study and informed them of their right to take breaks during the session and their ability to withdraw from the experiment at any time without providing a reason. Participants were then asked to read the information sheet and sign the consent form if they agreed to participate. Participants completed the vision test, the MoCA, and the sociodemographic and health questionnaire before playing SPACE. To prevent fatigue, participants were offered the option of a short break prior to starting the SPACE assessment. Each task in SPACE was explained verbally, and additional instructions were displayed within the user interface. Participants took 28 minutes on average to complete the tasks in SPACE. A detailed breakdown of the time required to complete each task under the various configurations is provided in Supplementary Table A2. After playing SPACE, participants filled out the System Usability Scale (SUS), User Experience Questionnaire (UEQ), NASA Task Load Index (NASA-TLX), Presence questionnaire, and a debriefing questionnaire. To collect gait data, participants walked a circuit for three minutes and then walked the circuit again while counting backwards for three minutes. The gait data were collected for future analyses and are not included in this paper.

### Analysis

We extracted the following performance variables from the tasks in SPACE. *Visuomotor training performance* was measured as the time (in seconds) required to complete the rotation, translation, circuit, and homing phases. *Path integration distance error* refers to the average distance between the participant’s final position and the target’s original position. Greater distances indicated larger errors. *Egocentric pointing error* was calculated as the average angular deviation (in degrees) between the participant’s estimate and the target landmark. *Mapping accuracy* was assessed using bidimensional regression (Tobler, 1965) to determine the degree of association (*R²*) between the real map of the environment and the map created by the participant. The A*ssociative memory score* was computed as the percentage of correct pairings between scanned elements and landmarks. *Perspective taking error* was measured as the average angular deviation (in degrees) between the participant’s estimate and the target landmark. In all analyses, we excluded the associative memory task as an outcome variable from our analysis because of ceiling effects, which limited variability and prevented the models from converging.

Before conducting inferential statistics, we verified whether our data violated the assumptions of the linear regression. Since some of the assumptions were violated, we used robust statistics to reduce the influence of outliers on the regression estimates by assigning them less weight in the model-fitting process (Field & Wilcox, 2017). We conducted three robust regression models with the MoCA score as the outcome variable. The first model included only age as a continuous predictor variable and gender as a dichotomous predictor variable. In addition to age and gender, the second model included the eight risk factors for dementia from the sociodemographic and health questionnaire (i.e., education, depression, anxiety, stress, alcohol intake, sleep duration, walking duration, and physical activity duration) as continuous predictor variables. Given that only four participants reported having no formal education, we combined participants with no formal education and those with a high school diploma into a single “lower education” category, contrasting it with participants who have a university-level education. The third model also included the tasks in SPACE (visuomotor, path integration, egocentric pointing, mapping, and perspective taking) as continuous predictors. We compared the first to the second model and the second to the third model using robust Wald tests and assessed the differences in fit in terms of changes in R^2^. Across all models, missing data were minimal. Models 1 and 2 each excluded one case due to a missing MoCA score, with Model 2 excluding one additional missing value for alcohol consumption. For Models 3, SPACE variables had limited missing values (Pointing: 2; Mapping: 2; Perspective: 10).

Next, we conducted a factor analysis with maximum likelihood extraction and varimax rotation on the MoCA and SPACE scores. Following Dwyer (Dwyer, 1937), we also used factor extension to evaluate the loadings of factors not included in the original analysis (i.e., age and gender). Finally, we generated age group and gender norms for each of the tasks in SPACE and visualised the data using continuous norming (Lenhard, Lenhard, Suggate, et al., 2018) across participant ages. All statistical analyses were performed using R Studio Version 2023.06.0+421 (R Studio PBC, Boston, MA, http://www.rstudio.com). Robust regressions and the Wald tests were conducted using the R packages *robustbase* (Maechler et al., 2023; Todorov & Filzmoser, 2009) and *WRS2 (Mair & Wilcox, 2020)*. We used the *psych* R package for the factor analysis (Revelle, 2007). Continuous norming was conducted using the *cNORM* R package (Lenhard, Lenhard, & Gary, 2018). The threshold for significance for all tests was set at α = .05.

## 3. RESULTS

Descriptive statistics for all predictor and outcome variables are listed in Table 2.

**Table 2.** Descriptive statistics for the dementia risk factors and the tasks in SPACE.

| Descriptive Statistics |  |  |  |
| --- | --- | --- | --- |
| Variable categorical | Level | Percentage |  |
| Gender | Female | 56.43 |  |
|  | Male | 43.57 |  |
| Education | High school | 27.49 |  |
|  | University | 72.51 |  |
| Variable continuous | Median | Mean | SD |
| MoCA | 27.00 | 26.68 | 2.33 |
| Age | 43.50 | 45.13 | 16.20 |
| Depression | 2.00 | 2.43 | 1.78 |
| Anxiety | 3.00 | 3.23 | 2.04 |
| Stress | 3.00 | 3.95 | 2.23 |
| Alcohol intake | 0.00 | 0.73 | 2.32 |
| Sleep | 7.00 | 6.75 | 0.90 |
| Walking | 7.00 | 9.95 | 9.61 |
| Physical activity | 2.00 | 2.80 | 2.86 |
| Visuomotor training | 241.29 | 246.14 | 37.15 |
| Path integration | 226.15 | 246.46 | 106.80 |
| Pointing | 66.45 | 65.33 | 20.32 |
| Mapping | 0.41 | 0.46 | 0.31 |
| Perspective taking | 20.64 | 28.70 | 23.12 |

The results of the regression models are presented in Table 3. The first robust regression model, including age and gender, significantly explained 10.7% of the variance in MoCA scores (χ*²*(2) = 38.278, *p* < 0.001). The results indicated that age has a significant negative effect on MoCA scores (β = -0.27, *p* < 0.001), and males performed significantly worse than females (β = -0.20, *p* = 0.033). The second model, including the individual risk factors as predictors in addition to age and gender, explained an additional 0.2% of the variance in MoCA scores. However, the second model did not significantly explain more variance than the first model (χ*²*(8) = 9.7729, *p* = 0.281). According to this second model, age (β = -0.24, *p* < 0.001) and gender (β = -0.24, *p* = 0.012) remained significant predictors. None of the individual risk factors significantly affected MoCA scores. The third model, including the scores from the spatial navigation tasks in SPACE, explained an additional 5.2% of the variance, which was a significant improvement over the second model (χ*²*(5) = 28.129, *p* < 0.001). According to this third model, age (β = -0.12, *p* = 0.044) and gender (β = -0.32, *p* < 0.001) remained significant predictors. Additionally, the pointing (β = -0.12, *p* = 0.031) and perspective taking (β = - 0.17, *p* = 0.007) tasks significantly predicted MoCA scores. We also tested whether SPACE tasks explained variance in MoCA scores beyond age and gender alone by fitting a model with only these predictors. Excluding dementia risk factors did not significantly reduce model fit (see Supplementary Information B1).

**Table 3.** Predictive models of MoCA scores using various risk factors for dementia and tasks from the SPACE assessment

|  | MoCA |  |  |
| --- | --- | --- | --- |
|  | Model 1 | Model 2 | Model 3 |
| Unstandardised estimates (std. Error) |  |  |  |
| (Intercept) | 28.874 (0.291)*** | 29.697 (1.149)*** | 32.205 (1.320)*** |
| Age | -0.038 (0.007)*** | -0.035 (0.007)*** | -0.017 (0.008)* |
| Gender<sub>[Male]</sub> | -0.460 (0.215)* | -0.561 (0.222)* | -0.742 (0.222)*** |
| Education<sub>[University]</sub> |  | 0.378 (0.240) | 0.201 (0.260) |
| Depression |  | 0.049 (0.072) | 0.034 (0.072) |
| Anxiety |  | 0.007 (0.090) | 0.033 (0.091) |
| Stress |  | -0.005 (0.081) | -0.019 (0.081) |
| N Alcohol |  | 0.041 (0.067) | 0.031 (0.083) |
| Sleep |  | -0.202 (0.136) | -0.168 (0.147) |
| Walking |  | -0.013 (0.014) | -0.015 (0.014) |
| Physical activity |  | 0.047 (0.034) | 0.032 (0.032) |
| Visuomotor training |  |  | -0.005 (0.003) |
| Path integration |  |  | -0.001 (0.001) |
| Pointing |  |  | -0.013 (0.006)* |
| Mapping |  |  | -0.687 (0.394) |
| Perspective taking |  |  | -0.017 (0.006)** |
| Num.Obs. | 341 | 340 | 330 |
| R<sup>2</sup> | 0.112 | 0.136 | 0.199 |
| R<sup>2</sup> Adj. | 0.107 | 0.110 | 0.161 |
| Residual Std. Error | 1.821 | 1.783 | 1.711 |

Note: **Model 1** (MoCA ~ Age + Gender); **Model 2** (MoCA ~ Age + Gender + Education + Depression + Anxiety + Stress + Alcohol intake + Sleep + Walking + Physical activity); **Model 3** (MoCA ~ Age + Gender + Education + Depression + Anxiety + Stress + Alcohol intake + Sleep + Walking + Physical activity + Visuospatial + Path integration + Pointing + Mapping + Perspective taking). \* $p < 0.05$ ; \*\* $p < 0.01$ ; \*\*\* $p < 0.001$ .

To confirm that the experimental conditions from Colombo and colleagues (2024) did not influence the results of Model 3, this model was reformulated as a linear mixed effects model with Condition included as a random factor. Including Condition as a random factor did not improve overall model fit and did not substantially change the pattern of significant effects in the models (see Supplementary Information C).

We also explored the extent to which the tasks in SPACE predicted different subdomains of the MoCA (see Supplementary Table D1 in Supplementary Information). We found that path integration (β = –.001, *p* < .001), pointing (β = –0.005, *p* = .026), mapping (β = -0.347, *p* = .017), and perspective taking (β = –0.009, *p* < .001) significantly predicted MoCA Visuospatial. Pointing (β = –0.003, *p* = .045) and perspective taking (β = –0.003, *p* = .028) also significantly predicted MoCA Abstraction. In addition, pointing significantly predicted MoCA Attention (β = –0.004, *p* = .004). Furthermore, perspective taking predicted MoCA Naming (β = –0.002, *p* = .011), MoCA Language (β = –0.01, *p* = .040), MoCA Delayed Recall (β = –0.008, *p* = .003), and MoCA Orientation (β = –0.001, *p* = .028).

The factor analysis included the variables age, gender, MoCA scores, and the tasks in the SPACE assessment. A correlation analysis revealed several significant relationships, suggesting potential underlying factors that could be extracted from the data (Figure 3).

**Figure 3.**
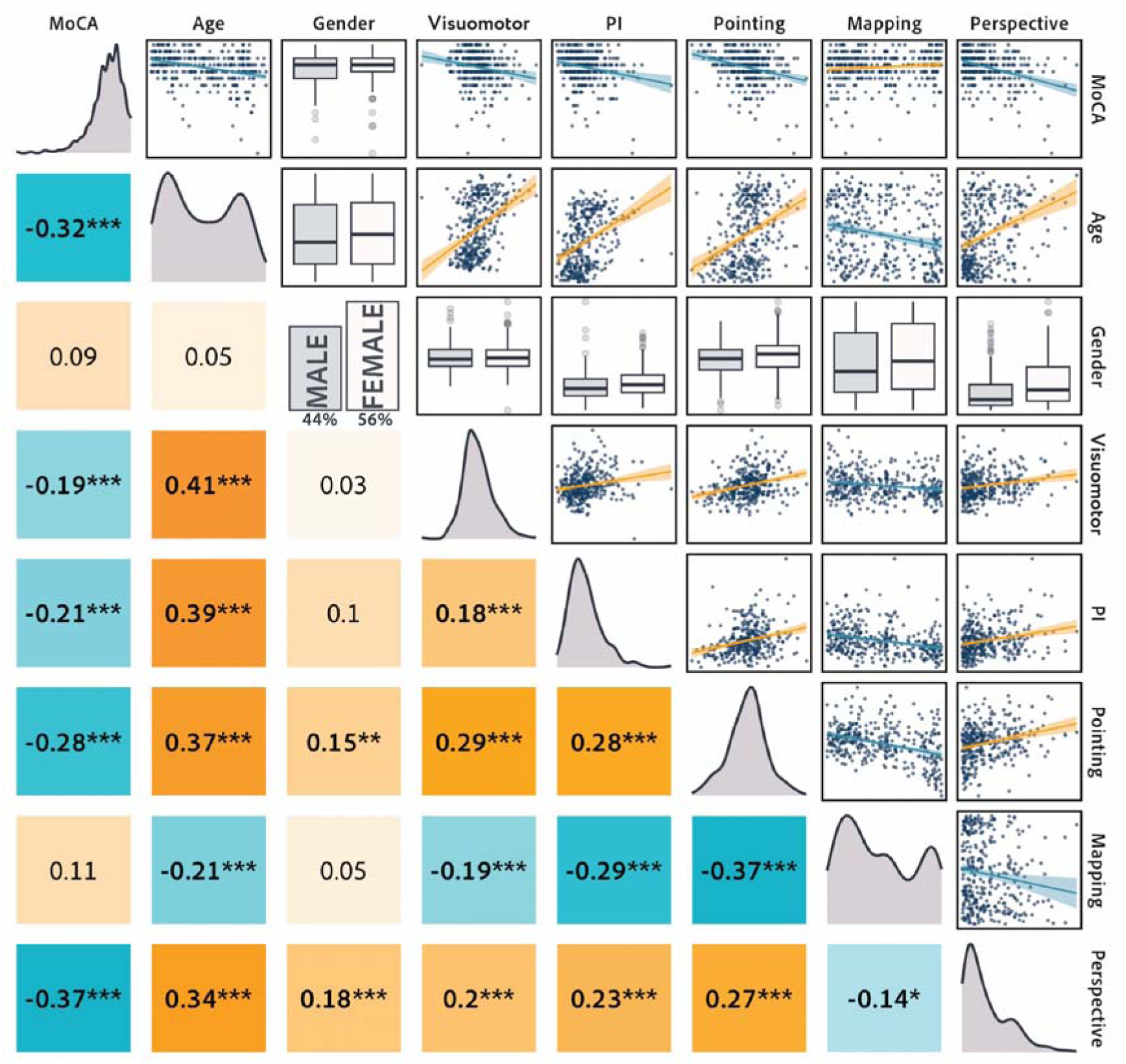
The correlation matrix presents Spearman’s correlation coefficients in the lower triangle, density plots along the diagonal illustrating data distributions, and scatterplots in the upper triangle showing the relationships between pairs of variables. Orange indicates positive correlations, and blue represents negative correlations. * p < 0.05; ** p < 0.01; *** p < 0.001.

Eigenvalues and a parallel analysis indicated the retention of two factors for the factor analysis (Figure 4). The eigenvalue for the first factor was 2.66, and the eigenvalue for the second factor was 1.10, indicating that these two factors together accounted for 23% of the total variance in the data (Figure 5). Specifically, ML1 and ML2 explained 8% and 15% of the total variance, respectively. ML1 had a moderate positive loading on visuomotor training (λ = 0.30), stronger positive loadings on pointing error (λ = 0.54) and path integration error (λ = 0.41) tasks, and a negative loading on the mapping accuracy (λ = -0.67). This factor appears to capture spatial and navigational abilities. ML2 had a strong positive loading on the perspective taking error (λ = 0.56) and a negative loading on MoCA scores (λ = -0.63), suggesting that this factor is related to cognitive and perceptual abilities (Figure 3). The standardised loadings for age and gender on ML1 and ML2 showed that age had moderate loadings on both factors (ML1:λ = 0.41, ML2: λ = 0.53), while gender had no impact (ML1: λ = 0.04, ML2: λ = 0.11). For the results of a factor analysis that included the risk factors, see Supplementary Table E1 in the Supplementary Information E.

**Figure 4.**
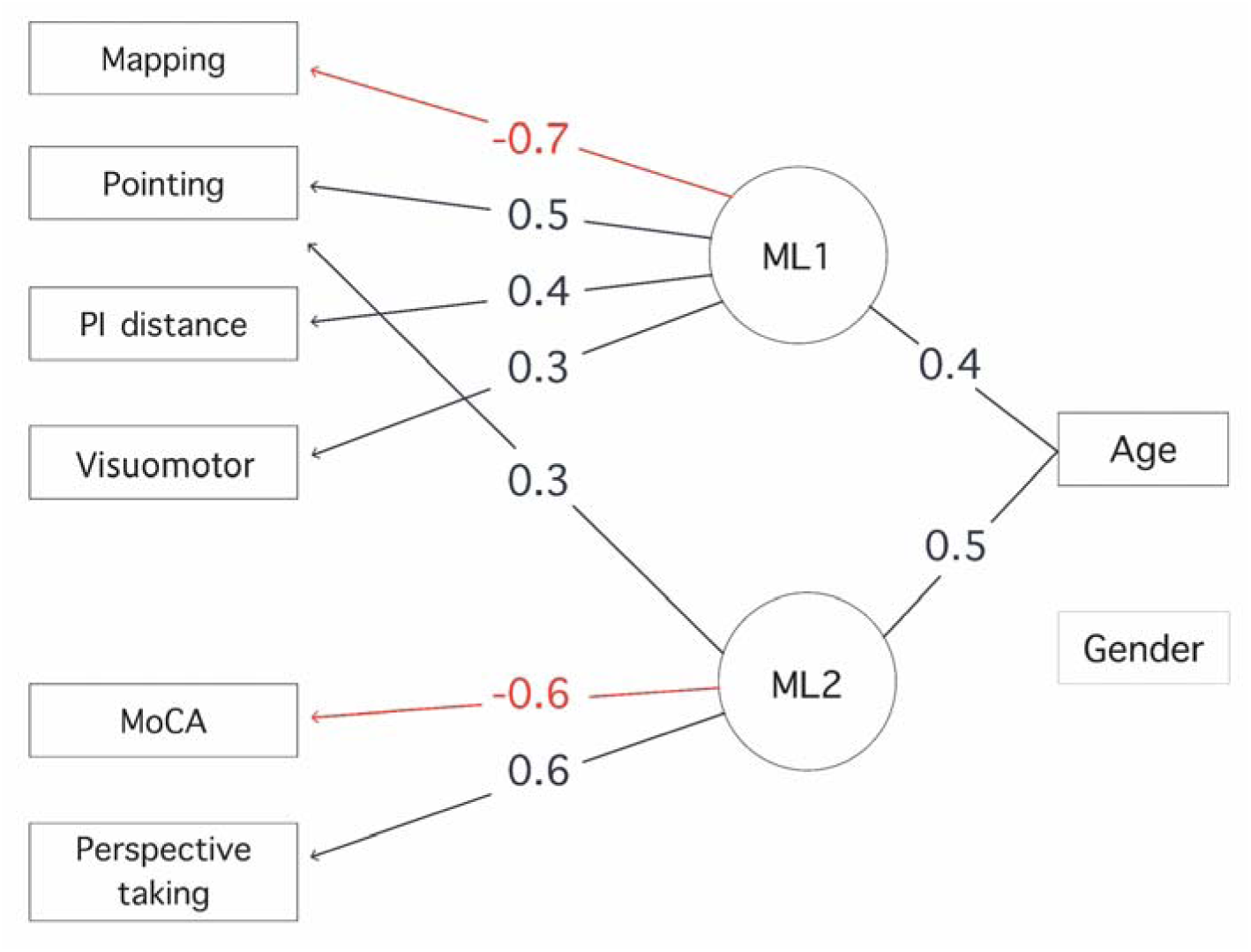
The diagram illustrates the results of the factor analysis for the SPACE tasks and MoCA scores, including the impact of age and gender extensions. Negative loadings are highlighted in red.

**Figure 5.**
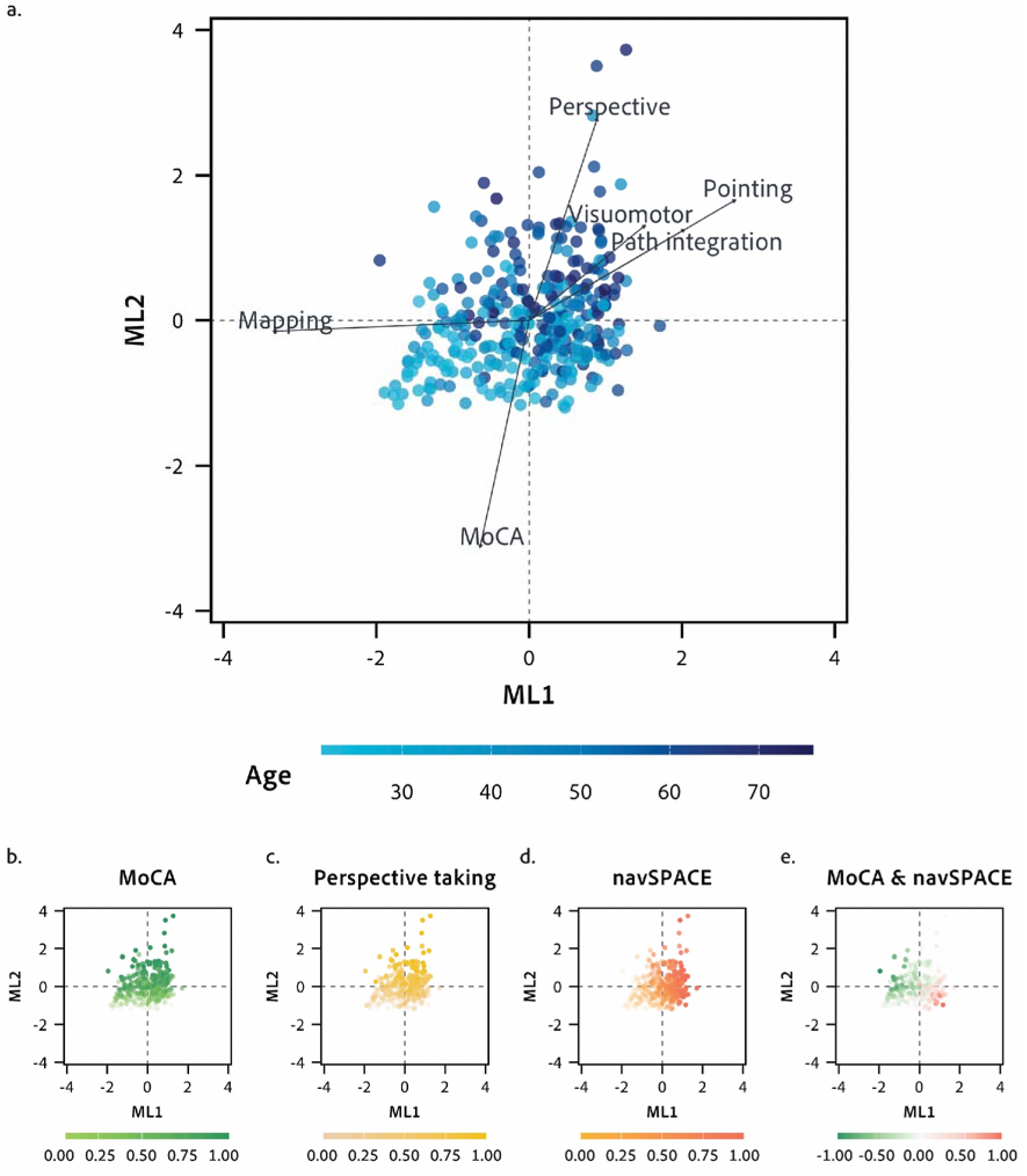
A series of scatter plots for ML1 and ML2, with each dot representing an individual participant. (a) The biplot displays loadings of MoCA and SPACE task scores on extracted factors (ML1 and ML2) and illustrates the relationships among variables. The length of the arrow for each variable represents the magnitude of the loading for that variable. The dots are coloured according to the participants’ ages. (b) The scatter plot of the factors ML1 and ML2 with dots coloured according to MoCA scores (reversed for visualisation) along a light green (low error) to dark green (high error) gradient. (c) The scatter plot of the factors ML1 and ML2 with dots coloured according to the errors on the perspective taking task along a light yellow (low error) to dark yellow (high error) gradient. (d) The scatter plot of the factors ML1 and ML2 with dots coloured according to errors from the navigation tasks in SPACE along a light orange (low error) to dark orange (high error) gradient. (e) The scatter plot of the factors ML1 and ML2 with dots coloured according to the difference between reversed MoCA scores and errors from the navigation tasks in SPACE along a gradient from green (difference favouring MoCA) to red (difference favouring the navigation tasks in SPACE). The visualisation demonstrates similar patterns for the MoCA and the perspective taking task. In addition, there are regions of the biplot representing participants performing worse on the navigation tasks in SPACE despite performing well on the MoCA. At the same time, some participants performed worse on the MoCA despite performing well on the navigation tasks in SPACE.

To facilitate the application of SPACE for the detection of cognitive impairment, we computed age and gender norms (Table 4) for each of the tasks in SPACE. Age norms are also listed by age group (i.e., 20-29, 30-39, 40-49, 50-59, and older than 60) and visualised using continuous norming in Figure 6.

**Figure 6.**
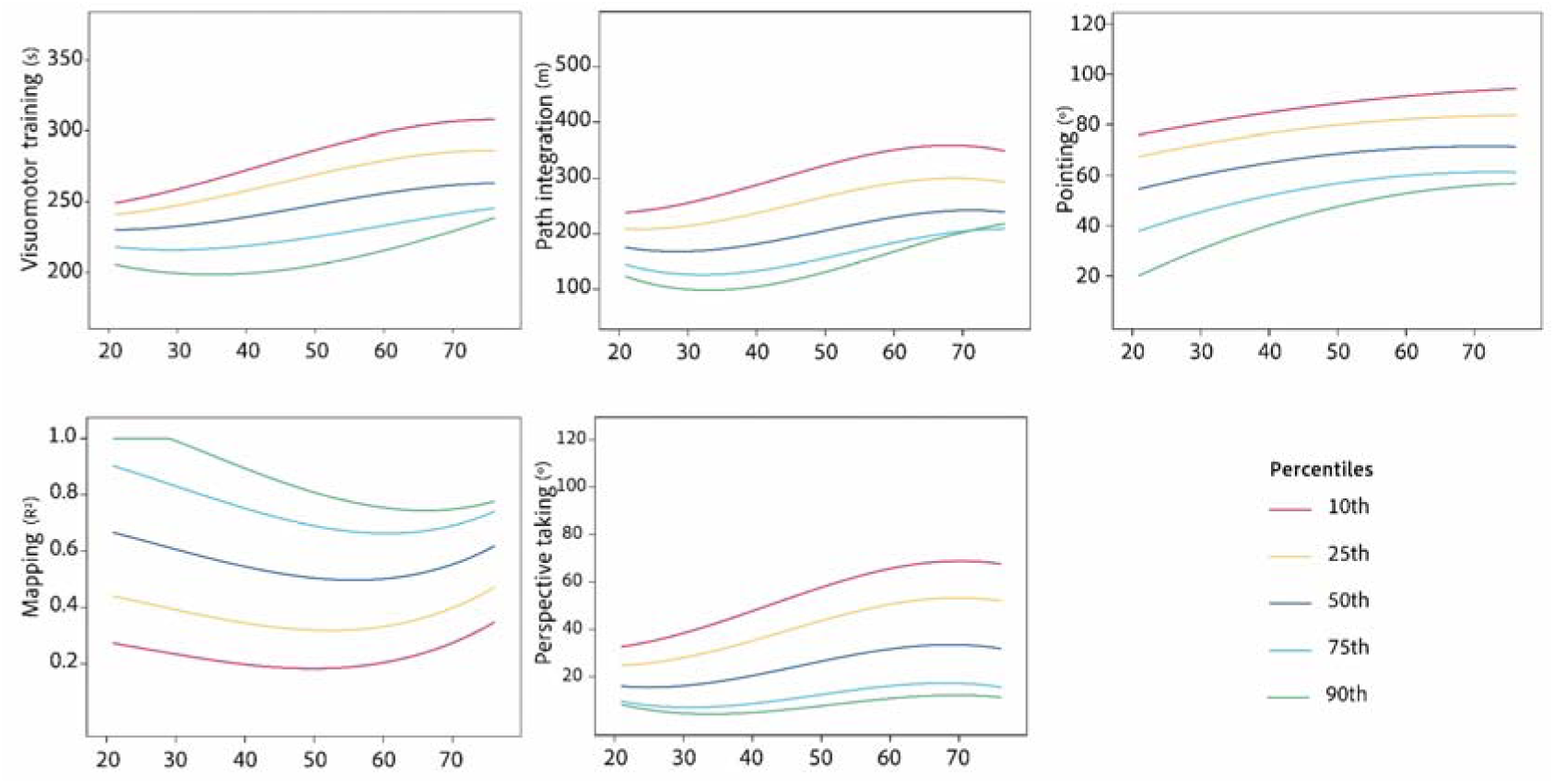
Age norms for the different tasks in SPACE. In the plots, error measures are used for visuomotor training, path integration, egocentric pointing, and perspective taking. The mapping task, however, assesses performance using an accuracy score.

**Table 4.** Normative data for age group and gender.

| Variable | Age | Gender | Mean | SD | 10th Percentile | 25th Percentile | 50th Percentile | 75th Percentile | 90th Percentile |
| --- | --- | --- | --- | --- | --- | --- | --- | --- | --- |
| Visuomotor training | 21-29 | Male | 223.02 | 27.62 | 187.97 | 204.86 | 221.79 | 233.29 | 258.38 |
|  |  | Female | 231.23 | 24.14 | 205.24 | 217.80 | 227.61 | 248.50 | 260.22 |
|  |  | Overall | 227.18 | 26.07 | 195.35 | 213.39 | 224.97 | 244.32 | 260.41 |
|  | 30-39 | Male | 236.18 | 24.44 | 208.60 | 222.92 | 236.15 | 248.79 | 264.41 |
|  |  | Female | 234.00 | 22.05 | 209.28 | 217.90 | 238.05 | 251.82 | 262.23 |
|  |  | Overall | 234.95 | 22.97 | 208.24 | 218.43 | 236.16 | 250.18 | 264.13 |
|  | 40-49 | Male | 237.26 | 32.37 | 195.07 | 220.85 | 234.60 | 260.95 | 282.28 |
|  |  | Female | 245.43 | 31.40 | 210.26 | 223.64 | 245.22 | 266.80 | 285.53 |
|  |  | Overall | 241.26 | 31.82 | 203.86 | 222.35 | 242.96 | 261.84 | 286.21 |
|  | 50-59 | Male | 251.09 | 28.54 | 221.58 | 231.31 | 241.57 | 266.98 | 291.71 |
|  |  | Female | 265.12 | 37.96 | 220.60 | 227.87 | 265.99 | 297.24 | 311.72 |
|  |  | Overall | 260.15 | 35.26 | 220.05 | 228.42 | 260.64 | 288.98 | 308.13 |
|  | 60+ | Male | 273.14 | 41.81 | 230.12 | 250.39 | 269.86 | 291.21 | 328.28 |
|  |  | Female | 261.51 | 42.78 | 218.58 | 233.96 | 248.96 | 291.14 | 321.88 |
|  |  | Overall | 266.33 | 42.55 | 220.07 | 239.79 | 258.38 | 291.53 | 324.45 |
| Path integration | 21-29 | Male | 189.62 | 67.10 | 96.06 | 152.43 | 182.89 | 226.77 | 267.45 |
|  |  | Female | 197.79 | 61.66 | 119.15 | 154.22 | 199.66 | 230.67 | 267.63 |
|  |  | Overall | 193.76 | 64.09 | 109.94 | 153.20 | 192.08 | 227.29 | 268.19 |
|  | 30-39 | Male | 211.51 | 97.70 | 110.41 | 130.93 | 210.73 | 259.99 | 334.77 |
|  |  | Female | 213.25 | 70.61 | 122.57 | 155.71 | 216.40 | 263.91 | 299.35 |
|  |  | Overall | 212.49 | 82.81 | 114.27 | 144.91 | 212.32 | 263.62 | 315.49 |
|  | 40-49 | Male | 197.38 | 70.17 | 124.10 | 153.40 | 172.20 | 235.61 | 305.64 |
|  |  | Female | 263.63 | 116.47 | 133.58 | 190.72 | 222.13 | 360.43 | 445.05 |
|  |  | Overall | 229.80 | 100.35 | 125.75 | 157.00 | 201.86 | 270.33 | 391.42 |
|  | 50-59 | Male | 220.41 | 81.35 | 154.05 | 191.27 | 202.29 | 249.72 | 301.16 |
|  |  | Female | 301.60 | 106.78 | 183.39 | 205.65 | 289.96 | 385.79 | 449.29 |
|  |  | Overall | 272.85 | 105.21 | 173.15 | 196.61 | 247.11 | 348.96 | 438.03 |
|  | 60+ | Male | 296.39 | 92.69 | 180.02 | 239.59 | 306.53 | 350.25 | 409.11 |
|  |  | Female | 301.27 | 105.95 | 175.85 | 230.78 | 287.24 | 348.80 | 441.17 |
|  |  | Overall | 299.25 | 100.17 | 179.26 | 232.78 | 291.64 | 350.42 | 427.16 |
| Pointing | 21-29 | Male | 52.95 | 20.80 | 24.66 | 44.49 | 51.76 | 67.07 | 75.54 |
|  |  | Female | 56.92 | 18.73 | 35.49 | 41.92 | 57.44 | 72.20 | 78.98 |
|  |  | Overall | 54.96 | 19.74 | 29.77 | 42.67 | 52.36 | 68.87 | 76.97 |
|  | 30-39 | Male | 59.71 | 18.43 | 30.97 | 50.19 | 65.14 | 72.14 | 81.57 |
|  |  | Female | 57.65 | 20.15 | 30.92 | 44.23 | 60.45 | 69.98 | 77.19 |
|  |  | Overall | 58.55 | 19.31 | 30.92 | 48.51 | 61.96 | 70.30 | 79.29 |
|  | 40-49 | Male | 65.67 | 18.49 | 43.00 | 55.89 | 66.03 | 81.66 | 87.22 |
|  |  | Female | 68.57 | 18.45 | 50.54 | 58.75 | 69.53 | 78.14 | 96.79 |
|  |  | Overall | 67.09 | 18.33 | 43.77 | 56.43 | 66.34 | 80.29 | 89.78 |
|  | 50-59 | Male | 64.49 | 14.61 | 47.51 | 56.76 | 65.92 | 74.92 | 77.30 |
|  |  | Female | 75.82 | 18.05 | 51.24 | 66.34 | 74.02 | 86.39 | 100.82 |
|  |  | Overall | 71.81 | 17.62 | 51.04 | 61.81 | 72.16 | 79.04 | 94.07 |
|  | 60+ | Male | 68.58 | 12.03 | 56.00 | 61.34 | 66.66 | 76.26 | 83.91 |
|  |  | Female | 77.97 | 15.86 | 57.91 | 67.02 | 75.71 | 86.55 | 101.21 |
|  |  | Overall | 74.14 | 15.08 | 56.69 | 64.31 | 73.21 | 82.43 | 96.99 |
| Mapping | 21-29 | Male | 0.51 | 0.33 | 0.94 | 0.88 | 0.50 | 0.20 | 0.11 |
|  |  | Female | 0.58 | 0.31 | 0.96 | 0.87 | 0.59 | 0.26 | 0.16 |
|  |  | Overall | 0.54 | 0.32 | 0.95 | 0.87 | 0.53 | 0.22 | 0.14 |
|  | 30-39 | Male | 0.48 | 0.33 | 0.95 | 0.83 | 0.49 | 0.20 | 0.10 |
|  |  | Female | 0.55 | 0.33 | 0.95 | 0.85 | 0.53 | 0.28 | 0.12 |
|  |  | Overall | 0.52 | 0.33 | 0.95 | 0.86 | 0.51 | 0.24 | 0.11 |
|  | 40-49 | Male | 0.52 | 0.36 | 0.95 | 0.89 | 0.56 | 0.20 | 0.10 |
|  |  | Female | 0.44 | 0.31 | 0.92 | 0.62 | 0.40 | 0.17 | 0.12 |
|  |  | Overall | 0.48 | 0.33 | 0.95 | 0.83 | 0.41 | 0.17 | 0.11 |
|  | 50-59 | Male | 0.39 | 0.30 | 0.88 | 0.51 | 0.28 | 0.19 | 0.10 |
|  |  | Female | 0.35 | 0.28 | 0.87 | 0.51 | 0.26 | 0.12 | 0.07 |
|  |  | Overall | 0.36 | 0.29 | 0.88 | 0.51 | 0.28 | 0.13 | 0.08 |
|  | 60+ | Male | 0.31 | 0.25 | 0.68 | 0.51 | 0.23 | 0.10 | 0.04 |
|  |  | Female | 0.43 | 0.29 | 0.89 | 0.62 | 0.34 | 0.18 | 0.08 |
|  |  | Overall | 0.38 | 0.28 | 0.82 | 0.56 | 0.29 | 0.15 | 0.06 |
| Perspective taking | 21-29 | Male | 13.30 | 10.26 | 4.54 | 5.85 | 10.41 | 18.92 | 23.91 |
|  |  | Female | 22.76 | 14.04 | 8.97 | 11.52 | 20.14 | 30.19 | 43.36 |
|  |  | Overall | 18.16 | 13.15 | 5.65 | 8.82 | 14.59 | 24.02 | 33.44 |
|  | 30-39 | Male | 22.74 | 20.15 | 5.26 | 8.53 | 12.96 | 35.88 | 51.62 |
|  |  | Female | 21.31 | 16.61 | 6.25 | 9.12 | 14.61 | 30.00 | 50.78 |
|  |  | Overall | 21.93 | 18.11 | 5.56 | 8.70 | 13.60 | 33.76 | 51.36 |
|  | 40-49 | Male | 23.89 | 25.98 | 6.45 | 8.90 | 13.05 | 22.72 | 59.91 |
|  |  | Female | 27.55 | 16.68 | 7.97 | 15.66 | 23.84 | 37.18 | 53.07 |
|  |  | Overall | 25.76 | 21.56 | 6.61 | 10.54 | 18.26 | 36.34 | 56.47 |
|  | 50-59 | Male | 25.27 | 19.07 | 5.82 | 10.31 | 22.91 | 32.02 | 49.12 |
|  |  | Female | 34.84 | 21.57 | 10.37 | 17.42 | 32.36 | 51.59 | 64.41 |
|  |  | Overall | 31.58 | 21.05 | 7.43 | 14.62 | 25.49 | 47.41 | 61.25 |
|  | 60+ | Male | 33.46 | 24.70 | 10.92 | 14.93 | 22.90 | 48.07 | 69.29 |
|  |  | Female | 44.30 | 26.55 | 12.87 | 24.94 | 39.26 | 62.08 | 73.83 |
|  |  | Overall | 39.80 | 26.21 | 11.53 | 19.38 | 33.07 | 60.17 | 73.00 |

## 4. DISCUSSION

This study investigates how dementia risk factors and performance on the spatial navigation tasks in SPACE predict MoCA scores as an indicator of cognitive impairment. The results of our regression analysis revealed that the pointing and perspective taking tasks in SPACE contributed to the prediction of MoCA scores beyond age and gender. Despite the established relationships between modifiable risk factors and cognitive impairment (Dale et al., 2018; Del Brutto et al., 2015; Livingston et al., 2024), none of the modifiable risk factors in our sample were significant predictors of MoCA scores. Our exploratory factor analysis further revealed that MoCA scores and performance on the perspective taking task in SPACE were associated with the same factor that was separate from the four navigation-related tasks in SPACE. In addition, we identified two clusters of participants who either performed well on the MoCA and poorly on the navigation tasks or well on the navigation tasks and poorly on the MoCA, suggesting that cognitive assessments may benefit from the combination of MoCA and navigation tasks. We argue that incorporating spatial navigation assessments into cognitive screening tests may improve their sensitivity and offer a more comprehensive and accurate evaluation of cognitive functioning.

A hallmark of MCI and AD is damage to the entorhinal cortex and hippocampus caused by the excessive accumulation of the β-amyloid peptides into neuritic plaques and an abnormal form of the protein tau into neurofibrillary tangles (Jack et al., 2010, 2024). Since structures in the Medial Temporal Lobe (MTL) are often associated with working memory and long-term declarative memory (Bird & Burgess, 2008; Eichenbaum, 2001; Squire, 1992), tasks such as the digit span task and delayed recall are typically used to detect cognitive impairment (Julayanont & Nasreddine, 2017). Indeed, the MoCA includes the digit span task and delayed recall, as well as visuospatial tasks such as trail-making and cube drawing (Julayanont & Nasreddine, 2017; Nasreddine et al., 2005). While both visuospatial tasks require patients to reconstruct small-scale spatial relations, these tasks do not involve the same scale and complexity of navigation skills associated with the MTL. Similar to how performance on declarative memory tasks is used to identify declarative memory impairment, performance on navigation tasks can contribute to the detection of the impairment of spatial skills that rely on the MTL but that are not yet assessed by the MoCA.

SPACE includes various navigation tasks such as visuomotor training, path integration, pointing, and mapping. There is now substantial evidence that performance on each of these individual tasks is associated with age (Lester et al., 2017; Stangl et al., 2020) and can discriminate with varying accuracy between healthy, MCI, and AD patients (deIpolyi et al., 2007; Howett et al., 2019; Mitolo et al., 2013; Segen et al., 2022; Tu et al., 2015). For example, Howett and colleagues (2019) tested healthy and MCI patients in an immersive path integration task and found that path integration error could discriminate between healthy participants and patients with MCI, especially for biomarker-positive patients (CSF amyloid-β and total tau). Notably, the ability of the path integration tasks to discriminate between biomarker-positive and biomarker-negative patients was significantly higher than the Trail Making Test-B and the Four Mountains Test. In addition, deIpolyi and colleagues (2007) found that, although MCI and mild AD patients could recognise landmarks along a learned route, these patients could not accurately identify landmark locations from a map or draw a map of the route. Similar to Howett and colleagues (2019), performance on a neuropsychological assessment, including the MMSE and measures of working memory and visuospatial memory, did not discriminate between patients with and without spatial impairments (deIpolyi et al., 2007). Comparable results were found in a virtual supermarket test in which participants were asked to orient to different goal locations after learning a route (2015). In that study, researchers found that the scores from this spatial orientation task discriminate between control, AD, and Frontotemporal Dementia participants. Together, this research suggests that spatial navigation assessments can complement traditional screenings for cognitive impairment and may support clinical decision-making.

While previous research focused on patients who had already been clinically diagnosed as cognitively impaired, we tested participants without a diagnosis and with a wide range of ages. We found that performance on the pointing and perspective taking tasks in SPACE predicted MoCA scores. Similarly, Tinella and colleagues (Tinella et al., 2022) found a significant correlation between MoCA scores and a perspective taking task of the same format (Kozhevnikov & Hegarty, 7 2001) for a large sample with a wide range of ages. Interestingly, this relationship was not found in other studies (Muffato et al., 2021; Muffato & De Beni, 2020) with a higher cutoff for MoCA scores (22 instead of 17), suggesting that perspective taking may be useful for discriminating between patients with different levels of cognitive impairment. Indeed, researchers have found that perspective taking tasks can discriminate between healthy and MCI participants (Laczó et al., 2021), healthy and AD participants (Chan et al., 2016; Laczó et al., 2021), and MCI and AD participants (Chan et al., 2016). Researchers have employed several variations of the perspective taking task, which have demonstrated good construct validity (Brucato et al., 2023). As part of SPACE, we introduce a variation of the perspective taking task that predicts MoCA scores and may be more scalable for broader deployment and unsupervised early screening (Tian et al., 2025).

Furthermore, we explored the factor structure underlying the MoCA and the tasks in SPACE, which revealed two distinct factors. The first factor included the MoCA and the perspective taking task, reinforcing the notion that the perspective taking task in SPACE could capture similar aspects of cognitive function assessed by the MoCA. The second factor combined the locomotion and wayfinding aspects of navigation tasks in SPACE (i.e., visuomotor training, path integration, pointing, and mapping) and may represent an overlooked dimension of cognitive functioning not captured by existing cognitive assessments. This factor structure is consistent with the findings of Meneghetti and colleagues (Meneghetti et al., 2021), who employed confirmatory factor analysis to derive the structure underlying visuospatial tasks and multiple wayfinding-related questionnaires. After finding that wayfinding inclinations (as derived from the questionnaires) underlie a separate factor from the visuospatial tasks, they showed that both factors predicted performance on navigation tasks in VR. Critically, the authors suggested that wayfinding inclinations predicted navigation recall performance because participants were asked to consider space at a larger scale.

Our results are supported by previous work that suggests a separation between small-scale and large-scale spatial abilities. For example, Meneghetti and colleagues (2014) found that small-scale perspective taking abilities cluster separately from large-scale navigation skills and exhibit stronger associations with general cognitive measures. Similarly, Malanchini and colleagues (2020) demonstrated that large-scale navigation abilities form a distinct, coherent factor from other spatial skills. In contrast, Hegarty and colleagues (Hegarty et al., 2006) showed that small-scale spatial tasks, such as mental rotation, are more strongly related to large-scale tasks in visual media than to learning in a real-world environment. However, they also found that a perspective taking task involving four objects was not related to tasks in either visual media or the real environment. Together, these findings suggest that perspective taking and large-scale navigation tasks represent different aspects of cognitive abilities. Indeed, while many of our participants performed either well or poorly on both SPACE and the MoCA, a substantial portion performed well on either SPACE or the MoCA. The present study provides evidence to support the claim that both types of spatial tasks may be used to complement existing screenings for cognitive impairment.

Visuomotor training was the only task in SPACE that did not predict the visuospatial subdomain of MoCA. This result may suggest that visuomotor training performance primarily reflects participants’ ability to locomote using the control interface and captures aspects of fine motor control, rate of learning, and reaction time, rather than large-scale spatial navigation. While these measures do not directly assess spatial abilities, they are nonetheless informative, as the rate of learning (Fernandez-Ballesteros et al., 2005; Wang et al., 2013) and motor performance (Li et al., 2024; Rudd et al., 2023) have been shown to be sensitive to subtle cognitive changes. For example, Fernández-Ballesteros and colleagues (2005) employed a dynamic assessment approach and found that individuals with MCI or mild AD exhibited reduced learning potential, as indicated by their diminished capacity to improve on cognitive tasks compared to that of healthy older adults. Recently, Li and colleagues (2024) employed a tablet-based “drawing and dragging” motor task, which achieved an accuracy of up to 85% in distinguishing individuals with MCI from healthy controls. In their study, older adults with MCI took significantly longer to switch between strokes, showed slower dragging speeds, and obtained lower overall task scores than cognitively healthy peers. In the present study, we may not have found a relationship between visuomotor training and MoCA scores because our sample was composed of healthy adults. However, our own research with clinical populations has shown that visuomotor training is a reliable predictor of MoCA scores (Colombo et al., 2025). Including visuomotor metrics alongside traditional navigation tasks may therefore provide additional insights into early variations in cognitive functioning.

Our factor analysis extension also revealed an effect of age on both factors (but no relationship with gender). According to previous research, spatial memory and related abilities tend to decline with age due to altered computations, functional deficits, and navigational impairments (Lester et al., 2017; Zancada-Menendez et al., 2016). As expected, our regression model revealed that older participants scored lower on the MoCA (Borland et al., 2017; Engedal et al., 2021; Gonçalves et al., 2023; Larouche et al., 2016; Santangelo et al., 2015; Thomann et al., 2018), and this difference may be more pronounced because of the wide age range in our sample. Our regression analysis also found that men had lower MoCA scores than women, aligning with prior research showing a female advantage in both older (Borland et al., 2017; Engedal et al., 2021; Thomann et al., 2018) and younger populations (Larouche et al., 2016), but see (Santangelo et al., 2015) for no differences and (Gonçalves et al., 2023) for a male advantage. Notably, none of the modifiable risk factors in our second model significantly predicted MoCA scores. Specifically, we did not observe a significant effect of education in our study, despite education often being associated with higher MoCA scores (Borland et al., 2017; Engedal et al., 2021; Gonçalves et al., 2023; Larouche et al., 2016; Santangelo et al., 2015; Thomann et al., 2018). These conflicting results may be attributable to the high education level of our sample, with only four participants without a high school diploma. The null effects for depression, anxiety, and stress may be attributable to a lack of sensitivity in our single-item scales for these conditions, although previous research suggests a high correspondence between single items and established measures such as the Depression, Anxiety and Stress Scale (Verster et al., 2021) and the Geriatric Depression Scale (McCormack et al., 2011). In our study, neither physical activity nor walking was associated with MoCA scores. While sustained physical activity has been found to protect against cognitive impairment (Iso-Markku et al., 2022), the results are mixed (Greendale et al., 2021; Wilson et al., 2002), and cognitive impairment can instead be the cause of a reduction in physical activity. Although previous studies link healthy lifestyles with a lower risk of dementia (Dhana et al., 2022), we found no association between alcohol consumption and MoCA scores in our sample. These results may be partially explained by the extremely low number of drinkers in our study. In more homogeneous and possibly healthier populations, these risk factors may have a reduced sensitivity to predict cognitive impairment.

The present study has at least three notable limitations. First, due to the inclusion of younger participants primarily from Singapore, we had less typical variation in several risk factors, including education. This sample prevented us from providing normative values for education level or geographic region. Second, we did not test participants longitudinally and do not know their eventual cognitive status in old age. Third, we focus on the results of a widely used cognitive assessment (i.e., the MoCA), which is not in itself a diagnostic tool. Clinical diagnoses would require a full neuropsychological exam, including tests for biomarkers of neurodegeneration. Despite these limitations, future work can benefit from incorporating SPACE into cognitive assessments, along with the normative data provided in the present study, for the early differentiation of healthy and pathological trajectories. Additionally, we plan to focus on more targeted groups by increasing recruitment of healthy and clinical middle-aged and older adults to strengthen the normative dataset and improve sensitivity to early cognitive changes over a longer period of time. Notably, these tests will include biomarkers as part of a diagnostic tool to further evaluate the utility of SPACE.

## 5. CONCLUSIONS

Digital assessments are becoming increasingly popular for assessing cognitive impairments (Berron, Olsson, et al., 2024; Liu et al., 2024; Meier et al., 2021; Thompson et al., 2023). SPACE differs from traditional cognitive assessments by providing spatial tasks in large and complex environments rather than small-scale tasks and questionnaires that focus solely on visuospatial skills, attention, and memory. Here, we show that SPACE predicts MoCA scores beyond traditional demographic and risk factors and has the potential to complement traditional tests for cognitive impairment.

## Data Availability

Data supporting this article may be available upon request and pending approval from the ethics committees in Singapore and ETH Zurich.

https://docs.google.com/document/d/1cy90i3if8AP1Dlenby-dhz7UZSPvbrP-SRYJIirB4ag/edit?usp=sharing

## ACKNOWLEDGMENTS

We would like to thank Tai Wei Lin Eunice, Martina Neudecker, Simona Margraf, and Sophia Niklaus for their assistance with data collection.

## 6. DECLARATIONS

### Ethics approval

Ethical approval for this study was granted by the Parkway Independent Ethics Committee (PIEC/2022/010) and the ETH Zurich Ethics Commission (EK 2021-N-193).

### Consent to participate

Written informed consent was obtained from all participants prior to their involvement in the study. All procedures adhered to the Declaration of Helsinki.

### Consent for publication

The research we conducted is not a case report. Only deidentified data from participants who signed the consent form is submitted for publication.

### Availability of data and materials

The data are not publicly available due to ethical constraints. Due to this limitation, the analysis code is illustrated using a synthetic dataset, which enables readers to verify the correctness of their implementation. The synthetic dataset can be found at the following link: https://osf.io/hyabv/?view_only=0eec51e300e14fdfba287a8e52a8f513.

### Code availability of data and materials

The code used for the analysis can be found at: https://osf.io/hyabv/?view_only=0eec51e300e14fdfba287a8e52a8f513.

### Authors’ contributions

**Giorgio Colombo:** Writing – review & editing, Writing – original draft, Visualisation, Software, Project administration, Methodology, Investigation, Formal analysis, Data curation, Conceptualisation. **Karolina Minta:** Writing – review & editing, Writing – original draft, Visualisation, Project administration, Methodology, Investigation, Formal analysis, Data curation, Conceptualisation. **Tyler Thrash:** Writing – review & editing, Writing – original draft, Formal analysis, Conceptualisation. **Jascha Grübel:** Writing – review & editing, Software, Formal analysis. **Jan Wiener:** Writing – review & editing. **Marios Avraamides:** Writing – review & editing. **William R. Taylor:** Writing – review & editing, Funding acquisition. **Christoph Hölscher:** Writing – review & editing. **Victor R. Schinazi:** Writing – review & editing, Writing – original draft, Supervision, Project administration, Funding acquisition, Formal analysis, Data curation, Conceptualisation.

