## Supplementary Information for "Beyond Traditional Assessments of Cognitive Status: Exploring the Potential of Spatial Navigation Tasks"

### Supplementary Materials

#### Supplementary Information A: Spatial layouts of the landmark configurations in the virtual environment

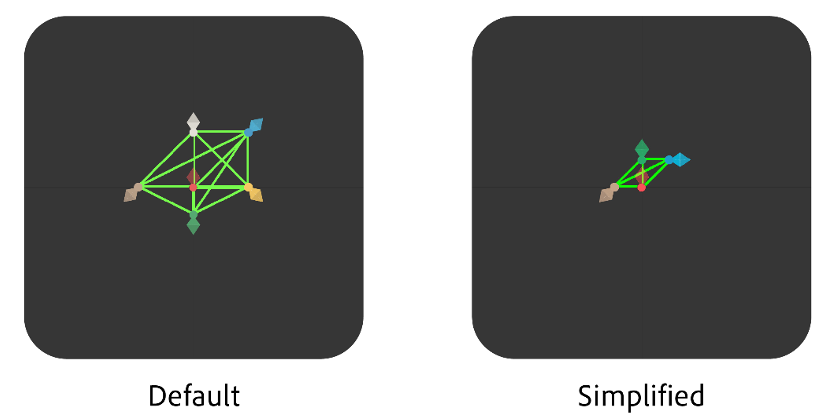

**Supplementary Figure A1.** Schematic representation of the Default or a Simplified configuration of landmarks in the virtual environment. The red rhombus represents the rocket's position. The other rhombuses represent the location of the other landmarks in the default and simplified configurations.

##### Proportion of participants across configurations and experimental conditions

This table summarises the proportion of participants who completed the SPACE assessment using either the Default or Simplified configuration of trials, and the corresponding experimental conditions. Overall, 64% of participants used the Default configuration (Tap, Anchor, and Widget), while 36% used the Simplified configuration. Because the Anchor control interface was used in every condition except Tap & Swipe, it was applied across both the Default configuration (Anchor and Widget conditions) and the Simplified configuration. As a result, 73% of participants used the Anchor interface, whereas 27% used Tap & Swipe. The table provides the full breakdown of participants by configuration and control condition.

**Supplementary Table A1.** Distribution of participants across SPACE configurations and control interface conditions.

| **Configuration** | **Condition** | **N (%)** |
| --- | --- | --- |
| **Default 220 (64%)** | Tap & Swipe control | 92 (27%) |
|  | Anchor control | 92 (27%) |
|  | UI Widget | 36 (10%) |
| **Simplified 122 (36%)** | Simplified trials | 122 (36%) |

##### Task completion and total SPACE duration across different configurations

To provide a detailed estimate of SPACE administration duration, we report mean completion times for each of the six component tasks (Visuomotor, Path Integration, Pointing, Mapping, Memory, and Perspective Taking) across the two usability configurations used in the study. On average, participants required approximately 36 minutes to complete SPACE in the Default condition and 21 minutes in the Simplified condition.

**Supplementary Table A2.** Mean task completion times (in seconds) for each SPACE task and total SPACE duration, reported separately for the Default and Simplified conditions. N refers to the number of participants per condition.

| **Summary of Task Times by Condition** | | | | | | | | |
| --- | --- | --- | --- | --- | --- | --- | --- | --- |
| Means (in seconds) | | | | | | | | |
| Configuration | Visuomotor | Path integration | Pointing | Mapping | Memory | Perspective taking | SPACE Time | N |
| Default | 243.43 | 1,061.11 | 355.30 | 82.13 | 41.27 | 448.12 | 2,194.07 | 220 |
| Simplified | 251.01 | 472.64 | 214.87 | 52.98 | 23.82 | 250.82 | 1,242.32 | 122 |

#### Supplementary Information B: Regression of age, gender and SPACE predicting MoCA scores excluding the risk factors

To examine whether the SPACE tasks provide added explanatory value relative to age and gender, irrespective of dementia risk factors, we compared nested models using robust Wald tests. Adding SPACE measures to the risk-factor model significantly improved model fit (Model 2 → Model 3). Importantly, removing the risk factors from the full model did not significantly reduce model performance (Model 3 → Model 4), χ²(8) = 6.07, *p* = .639. This result indicates that the dementia risk factors do not meaningfully contribute to variance in MoCA scores once the SPACE tasks are included. Because the risk factor model (Model 2) and the SPACE-only model (Model 4) are not nested, formal model comparison via ANOVA is not applicable. Instead, we compared the models using pseudo-R² and RMSE. The SPACE-only model showed higher explanatory power (pseudo-R² = 0.167) and lower prediction error (RMSE = 2.06) than the risk-factor model (pseudo-R² = 0.110; RMSE = 2.17).

**Supplementary Table B1.** Robust regression of MoCA scores, including and excluding dementia risk factors.

| Regression predicting MoCA | | | | |
| --- | --- | --- | --- | --- |
|  | Model 1 | Model 2 | Model 3 | Model 4 |
| (Intercept) | **28.874 (0.291)***** | **29.697 (1.149)***** | **32.205 (1.320)***** | **31.336 (0.834)***** |
| Age | **-0.038 (0.007)***** | **-0.035 (0.007)***** | **-0.017 (0.008)*** | **-0.020 (0.008)*** |
| Gender _[Male]_ | **-0.460 (0.215)*** | **-0.561 (0.222)*** | **-0.742 (0.222)***** | **-0.680 (0.216)**** |
| Education _[University]_ |  | 0.378 (0.240) | 0.201 (0.260) |  |
| Depression |  | 0.049 (0.072) | 0.034 (0.072) |  |
| Anxiety |  | 0.007 (0.090) | 0.033 (0.091) |  |
| Stress |  | -0.005 (0.081) | -0.019 (0.081) |  |
| N Alcohol |  | 0.041 (0.067) | 0.031 (0.083) |  |
| Sleep |  | -0.202 (0.136 | -0.168 (0.147) |  |
| Walking |  | -0.013 (0.014) | -0.015 (0.014) |  |
| Physical activity |  | 0.047 (0.034) | 0.032 (0.032) |  |
| Visuomotor training |  |  | -0.005 (0.003) | -0.005 (0.003) |
| Path integration |  |  | -0.001 (0.001) | -0.001 (0.001) |
| Pointing |  |  | -0.013 (0.006)* | **-0.014 (0.006)*** |
| Mapping |  |  | -0.687 (0.394) | -0.656 (0.375) |
| Perspective taking |  |  | -0.017 (0.006)** | **-0.019 (0.006)**** |
| Num.Obs. | 341 | 340 | 330 | 331 |
| R2 | 0.112 | 0.136 | 0.199 | 0.185 |
| R2 Adj. | 0.107 | 0.110 | 0.161 | 0.167 |
| RMSE | 2.21 | 2.17 | 2.04 | 2.06 |
| Note: Model 1: MoCA ~ Age + Gender; Model 2: MoCA ~ Age + Gender + Education + Depression + Anxiety + Stress + Alcohol + Sleep + Walking + Physical Activity + Visuomotor Training; Model 3: MoCA ~ Age + Gender + Education + Depression + Anxiety + Stress + Alcohol + Sleep + Walking + Physical Activity + Visuomotor Training + Path Integration + Pointing + Mapping + Perspective Taking; Model 4: MoCA ~ Path Integration + Pointing + Mapping + Perspective Taking. p < 0.1, * p < 0.05, ** p < 0.01, *** p < 0.001. | | | | |

#### Supplementary Information C: Controlling for Experimental Conditions

The study sample in the current manuscript partially overlaps with our previous usability publication on SPACE, but addresses a different research question. Whereas the prior work compared four usability conditions to evaluate interface design and user experience, the present study focuses on individual differences in spatial ability and their relationship with demographic, cognitive, and dementia-relevant risk factors. To maximise statistical power, participants from all usability configurations were pooled and supplemented with 87 additional recruits (total N = 348). Because participants completed the SPACE under different interface conditions, we verified whether these conditions influenced the prediction of MoCA scores by fitting a linear mixed-effects model that included Condition as a random factor. The model fit did not improve relative to the corresponding fixed-effects model (LRT: χ² = 0.648, p = 0.421), indicating that the interface condition did not contribute meaningfully to MoCA variance. Importantly, the same predictors (gender, pointing, and perspective taking) remained significant in both models. Supplementary Table C1 reports the distribution of participants across interface configurations and control conditions.

**Supplementary Table C1.** Linear mixed-effects model examining whether interface condition influenced MoCA scores when controlling for demographic variables, risk factors, and SPACE task performance. Condition was included as a random effect and did not significantly improve model fit. Significant predictors (age, gender, pointing, and perspective taking) remained consistent across models.

|  | **MoCA** | | | | | | | |
| --- | --- | --- | --- | --- | --- | --- | --- | --- |
| Predictors | Estimates | std. Error | std. Beta | standardised std. Error | CI | standardised CI | Statistic | p |
| (Intercept) | 33.00 | 1.47 | 0.09 | 0.12 | 30.11 – 35.89 | -0.15 – 0.33 | 22.47 | **<0.001** |
| Age | -0.02 | 0.01 | -0.11 | 0.07 | -0.03 – 0.00 | -0.24 – 0.02 | -1.72 | 0.086 |
| Gender _[Male]_ | -0.97 | 0.24 | -0.42 | 0.10 | -1.45 – -0.49 | -0.62 – -0.21 | -4.01 | **<0.001** |
| Education _[University]_ | 0.26 | 0.27 | 0.11 | 0.12 | -0.29 – 0.80 | -0.12 – 0.34 | 0.93 | 0.354 |
| Depression | 0.01 | 0.10 | 0.01 | 0.07 | -0.18 – 0.20 | -0.14 – 0.15 | 0.11 | 0.911 |
| Anxiety | -0.00 | 0.10 | -0.00 | 0.09 | -0.20 – 0.19 | -0.18 – 0.17 | -0.05 | 0.963 |
| Stress | 0.02 | 0.08 | 0.02 | 0.08 | -0.14 – 0.19 | -0.14 – 0.18 | 0.28 | 0.779 |
| N Alcohol | 0.04 | 0.05 | 0.04 | 0.05 | -0.06 – 0.14 | -0.06 – 0.14 | 0.81 | 0.420 |
| Sleep | -0.25 | 0.13 | -0.09 | 0.05 | -0.51 – 0.02 | -0.19 – 0.01 | -1.85 | 0.065 |
| Walking | -0.02 | 0.01 | -0.10 | 0.05 | -0.05 – 0.00 | -0.20 – 0.00 | -1.94 | 0.053 |
| Physical activity | 0.04 | 0.04 | 0.04 | 0.05 | -0.05 – 0.12 | -0.06 – 0.14 | 0.85 | 0.397 |
| Visuospatial | -0.00 | 0.00 | -0.03 | 0.06 | -0.01 – 0.01 | -0.14 – 0.08 | -0.57 | 0.569 |
| PI distance | -0.00 | 0.00 | -0.08 | 0.06 | -0.00 – 0.00 | -0.20 – 0.03 | -1.39 | 0.164 |
| Pointing | -0.02 | 0.01 | -0.21 | 0.06 | -0.04 – -0.01 | -0.33 – -0.09 | -3.55 | **<0.001** |
| Mapping | -0.70 | 0.43 | -0.10 | 0.06 | -1.54 – 0.13 | -0.21 – 0.02 | -1.65 | 0.099 |
| Perspective | -0.03 | 0.01 | -0.28 | 0.06 | -0.04 – -0.02 | -0.39 – -0.17 | -5.07 | **<0.001** |
| **Random Effects** | | | | | | | | |
| σ^2^ | 4.13 | | | | | | | |
| τ_00_ _Condition_ | 0.09 | | | | | | | |
| ICC | 0.02 | | | | | | | |
| N _Condition_ | 4 | | | | | | | |
| LRT χ² / p value | 0.648 / 0.421 | | | | | | | |
| Observations | 330 | | | | | | | |
| Marginal R^2^ /  Conditional R^2^ | 0.260 / 0.275 | | | | | | | |

#### Supplementary Information D: Associations Between SPACE Tasks and MoCA Subdomains

To provide a more fine-grained examination of the relationship between SPACE performance and general cognition, we conducted linear regressions with each MoCA subdomain (Visuospatial, Naming, Attention, Language, Abstraction, Delayed Recall, Orientation) as the outcome variable and SPACE task performance as predictors while controlling for age and gender. While the total MoCA score captures broad cognitive functioning, analysing subdomains allows the identification of domain-specific associations. Results (Supplementary Table D1) indicate that MoCA Visuospatial performance was significantly predicted by nearly all SPACE tasks, consistent with the spatial demands of the task. Perspective taking emerged as the most consistent predictor across multiple subdomains, including Naming, Language, Delayed Recall, and Orientation, suggesting it engages broader cognitive processes such as executive function and spatial transformation. Pointing showed more selective associations with Attention and Abstraction.

**Supplementary Table D1.** Linear regression models examining the association between SPACE task performance and individual MoCA subdomains. The table reports unstandardized regression coefficients (β), standard errors (SE), t-values, and p-values for each predictor across the MoCA subdomains (Visuospatial, Naming, Attention, Language, Abstraction, Delayed Recall, and Orientation). Predictors include demographic variables (age and gender) and SPACE tasks (visuospatial training, path integration, pointing, mapping, and perspective-taking). The model fit (R²) and sample size are provided for each domain.

| **MoCA subdomains** | | | | | | |
| --- | --- | --- | --- | --- | --- | --- |
|  | Predictor | β | (SE) | t value | p value | R² / N |
| **Visuospatial** | Intercept | 5.399 | (0.338) | 15.96 | <.001*** | 0.125 / 331 |
|  | Age | 0.005 | (0.003) | 1.46 | 0.145 |  |
|  | Gender | -0.214 | (0.086) | -2.50 | 0.013* |  |
|  | Visuomotor training | 0.000 | (0.001) | 0.20 | 0.841 |  |
|  | Path integration | -0.001 | (0.000) | -3.34 | <.001*** |  |
|  | Pointing | -0.005 | (0.002) | -2.24 | 0.026* |  |
|  | Mapping | -0.347 | (0.145) | -2.39 | 0.017* |  |
|  | Perspective | -0.009 | (0.002) | -4.45 | <.001*** |  |
| **Naming** | Intercept | 3.066 | (0.141) | 21.77 | <.001*** | 0.047 / 331 |
|  | Age | -0.001 | (0.001) | -0.88 | 0.378 |  |
|  | Gender | -0.019 | (0.036) | -0.53 | 0.595 |  |
|  | Visuomotor training | 0.000 | (0.001) | 0.58 | 0.560 |  |
|  | Path integration | -0.000 | (0.000) | -0.39 | 0.697 |  |
|  | Pointing | -0.001 | (0.001) | -1.29 | 0.197 |  |
|  | Mapping | -0.009 | (0.060) | -0.15 | 0.880 |  |
|  | Perspective | -0.002 | (0.001) | -2.56 | 0.011* |  |
| **Attention** | Intercept | 6.421 | (0.199) | 32.24 | <.001*** | 0.067 / 331 |
|  | Age | 0.000 | (0.002) | 0.02 | 0.983 |  |
|  | Gender | -0.045 | (0.050) | -0.90 | 0.368 |  |
|  | Visuomotor training | -0.001 | (0.001) | -1.01 | 0.311 |  |
|  | Path integration | -0.000 | (0.000) | -0.81 | 0.418 |  |
|  | Pointing | -0.004 | (0.001) | -2.86 | 0.004** |  |
|  | Mapping | -0.066 | (0.086) | -0.77 | 0.444 |  |
|  | Perspective | -0.002 | (0.001) | -1.82 | 0.069 |  |
| **Language** | Intercept | 2.686 | (0.376) | 7.14 | <.001*** | 0.145 / 331 |
|  | Age | -0.017 | (0.004) | -4.86 | <.001*** |  |
|  | Gender | -0.260 | (0.095) | -2.73 | 0.007** |  |
|  | Visuomotor training | 0.000 | (0.001) | 0.34 | 0.734 |  |
|  | Path integration | 0.000 | (0.000) | 0.82 | 0.415 |  |
|  | Pointing | -0.002 | (0.003) | -0.93 | 0.351 |  |
|  | Mapping | -0.102 | (0.162) | -0.63 | 0.529 |  |
|  | Perspective | -0.005 | (0.002) | -2.38 | 0.018* |  |
| **Abstraction** | Intercept | 2.042 | (0.240) | 8.50 | <.001*** | 0.041 / 331 |
|  | Age | 0.003 | (0.002) | 1.20 | 0.230 |  |
|  | Gender | -0.010 | (0.061) | -0.17 | 0.865 |  |
|  | Visuomotor training | -0.000 | (0.001) | -0.01 | 0.994 |  |
|  | Path integration | -0.000 | (0.000) | -1.38 | 0.169 |  |
|  | Pointing | -0.003 | (0.002) | -2.01 | 0.045* |  |
|  | Mapping | -0.115 | (0.103) | -1.11 | 0.266 |  |
|  | Perspective | -0.003 | (0.001) | -2.21 | 0.028* |  |
| **Delayed recall** | Intercept | 5.644 | (0.461) | 12.24 | <.001*** | 0.125 / 331 |
|  | Age | -0.010 | (0.004) | -2.35 | 0.019* |  |
|  | Gender | -0.270 | (0.117) | -2.31 | 0.021* |  |
|  | Visuomotor training | -0.001 | (0.002) | -0.80 | 0.422 |  |
|  | Path integration | -0.000 | (0.001) | -0.06 | 0.948 |  |
|  | Pointing | -0.005 | (0.003) | -1.66 | 0.098 |  |
|  | Mapping | 0.086 | (0.198) | 0.43 | 0.665 |  |
|  | Perspective | -0.008 | (0.003) | -3.04 | 0.003** |  |
| **Orientation** | Intercept | 6.004 | (0.081) | 74.14 | <.001*** | 0.028 / 331 |
|  | Age | -0.000 | (0.001) | -0.58 | 0.565 |  |
|  | Gender | -0.011 | (0.021) | -0.54 | 0.589 |  |
|  | Visuomotor training | 0.000 | (0.000) | 0.02 | 0.987 |  |
|  | Path integration | 0.000 | (0.000) | 1.52 | 0.130 |  |
|  | Pointing | -0.000 | (0.001) | -0.11 | 0.911 |  |
|  | Mapping | -0.042 | (0.035) | -1.20 | 0.233 |  |
|  | Perspective | -0.001 | (0.000) | -2.21 | 0.028* |  |

#### Supplementary Information E: Factor analysis of SPACE including dementia risk factors

An exploratory factor analysis with varimax rotation was conducted to examine the underlying structure of cognitive (MoCA), spatial (SPACE tasks), and modifiable dementia risk factors (depression, anxiety, stress, alcohol use, sleep, walking, physical activity). Eigenvalues and parallel analysis supported the retention of three factors (ML1, ML2, ML3), explaining 32.7% of the total variance (ML1 = 16.4%, ML2 = 8.5%, ML3 = 7.8%). ML1 loaded strongly on depression (λ = 0.78), anxiety (λ = 0.93), and stress (λ = 0.80), representing a mental health factor. ML2 showed a moderate negative loading on MoCA (λ = -0.62) and a positive loading on perspective taking (λ = 0.55), reflecting cognitive and perceptual skills. ML3 showed positive loadings on mapping (λ = 0.54) and negative loadings on path integration (λ = -0.38) and pointing (λ = -0.58), capturing spatial and navigation skills. Standardised loadings of age and gender indicated that age had a minor influence on ML1 (λ = -0.24) and a moderate influence on ML2 (λ = 0.50) and ML3 (λ = -0.36), while gender had negligible effects on all factors. Importantly, the modifiable risk factors did not load significantly onto ML2 or ML3, suggesting that the SPACE measures and MoCA capture variance independently of these risk factors.

**Supplementary Table E1.** Exploratory factor analysis of cognitive, spatial, and dementia risk factors.

| **Variable** | **ML1** | **ML2** | **ML3** |
| --- | --- | --- | --- |
| MoCA | — | **–0.62** | — |
| Visuomotor training | — | 0.32 | –0.28 |
| Path integration | — | 0.28 | **–0.38** |
| Pointing | — | 0.35 | **–0.58** |
| Mapping | — | –0.12 | **0.54** |
| Perspective Taking | — | **0.55** | –0.15 |
| Depression | **0.78** | — | 0.11 |
| Anxiety | **0.93** | — | 0.15 |
| Stress | **0.80** | –0.21 | — |
| N Alcohol | — | –0.11 | — |
| Sleep | — | — | — |
| Walking | — | 0.18 | 0.22 |
| Physical Activity | — | 0.22 | — |
